# Transfer learning with randomized controlled trial data for postprandial glucose prediction

**DOI:** 10.1101/2024.01.28.24301902

**Authors:** Shinji Hotta, Mikko Kytö, Saila Koivusalo, Seppo Heinonen, Pekka Marttinen

**Affiliations:** Department of Computer Science, Aalto University, Espoo, Finland; Fujitsu Limited, Kawasaki, Japan; IT Management, Helsinki University Hospital, Helsinki, Finland; Department of Computer Science, University of Helsinki, Helsinki, Finland; Shared Group Services, Helsinki University Hospital, University of Helsinki, Helsinki, Finland; Department of Obstetrics and Gynecology, Helsinki University Hospital, University of Helsinki, Helsinki, Finland

## Abstract

In recent years, numerous methods have been introduced to predict glucose levels using machine-learning techniques on patients’ daily behavioral and continuous glucose data. Nevertheless, a definitive consensus remains elusive regarding modeling the combined effects of diet and exercise for optimal glucose prediction. A notable challenge is the propensity for observational patient datasets from uncontrolled environments to overfit due to skewed feature distributions of target behaviors; for instance, diabetic patients seldom engage in high-intensity exercise post-meal. In this study, we introduce a unique Bayesian transfer learning framework using randomized controlled trial (RCT) data, primarily targeting postprandial glucose prediction. Initially, we gathered balanced training data from RCTs on healthy participants by randomizing behavioral conditions. Subsequently, we pretrained the model’s parameter distribution using RCT data from the healthy cohort. This pretrained distribution was then adjusted, transferred, and utilized to determine the model parameters for each patient. Our framework’s efficacy was appraised using data from 68 gestational diabetes mellitus patients in uncontrolled settings. The evaluation underscored the enhanced performance attained through our framework. Furthermore, when modeling the joint impact of diet and exercise, the synergetic model proved more precise than its additive counterpart.

## Introduction

The global incidence of diabetes is on the rise, accompanied by escalating severity. This progression entails detrimental ramifications including compromised quality of life (QoL), multifarious complications, and costly surgical treatment. Projections indicate a staggering $2.5 trillion global expenditure on diabetes-related medical costs by 2030 [1]. In light of this, there is an imperative to curtail these expenses while ameliorating QoL by proactively mitigating diabetes severity.

Recent national clinical guidelines [34] underscore a fundamental tenet of preventive intervention: the maintenance of blood glucose levels within the normative spectrum. A pivotal approach to achieving this control involves adopting a balanced lifestyle encompassing dietary measures, physical activity, and insulin therapy. Technological strides in continuous glucose monitoring (CGM) apparatus have empowered patients in managing glucose levels via mobile applications in the comfort of their homes, facilitating self-care [2,3]. However, mastering optimal behavioral adjustments for maintaining normoglycemia remains a challenge for patients [4]. Hence, a personalized framework is imperative, one that tailors recommendations for optimal individual behaviors, thereby ensuring the trajectory of future blood glucose levels aligns with the norm. This ambition necessitates the precise anticipation of how behavioral modifications will influence forthcoming blood glucose dynamics.

To date, various data-driven techniques for predicting glucose, incorporating behavioral factors, have emerged [5,6]. Prior literature primarily focused on dietary and exercise facets, employing time-series machine learning models like the autoregressive model [7] and long short-term memory (LSTM) [8] for glucose forecasting. Independent positive impacts on predictive accuracy have been demonstrated for dietary and exercise factors [6]. Yet, the synergy between these factors in achieving optimal prediction remains understudied. Recent clinical investigations [9–13] have illuminated that strategic synchronization of diet and exercise, such as moderate postprandial exercise, holds potential for further glucose reduction across diabetes profiles. However, the translation of these discoveries into predictive glucose modeling has remained uncharted.

When considering the amalgamation of multiple behaviors, a critical hurdle is sidestepping overfitting in learning from an imbalanced patient dataset collected in unconstrained settings. Taking the instance of diet and exercise integration, the frequency of intermediate-level post-meal exercise tends to be notably lower than that of minimal or no post-meal exercise among gestational diabetes mellitus (GDM) patients leading their daily lives [38]. Consequently, an accurate estimation of the combined impact of postprandial exercise and diet becomes challenging due to sparse data on high-intensity postprandial exercise.

In recent years, an innovative solution has emerged to tackle data imbalance concerns by harnessing extensive patient data through transfer learning techniques [8,14–16]. This strategy thrives when the feature distributions of other patient data exhibit a range of values. Yet, when feature distribution across all patients is markedly imbalanced, the risk of extracting and transferring inaccurate insights escalates. Our preliminary analysis indeed revealed a pronounced imbalance in the feature distribution of postprandial exercises among 68 gestational diabetes mellitus (GDM) patients observed over 18 days (Fig 7(a)).

This paper introduces a novel learning framework (Fig 1) that synergizes transfer learning with supplementary intervention data from a randomized controlled trial (RCT), harmonizing the distribution of behavioral features. This harmony is instrumental in predictive modeling of postprandial glucose involving dietary and exercise variables. The framework commences with an RCT, where behavioral conditions are randomized for a healthy cohort, amassing data with a balanced distribution. Subsequently, Bayesian parameter learning is executed on the prediction model utilizing the RCT data, yielding a dependable parameter distribution. Ultimately, this pre-trained distribution is judiciously rescaled and employed as a prior for each patient’s parameter learning utilizing observational data from real-world scenarios. This ensures a robust knowledge transfer from the RCT domain, curtailing overfitting risks inherent in imbalanced patient data. Empirical validation underscores the efficacy of this framework, as evidenced by enhanced prediction performance in postprandial glucose prognosis using an authentic GDM patient dataset.

**Fig 1.**
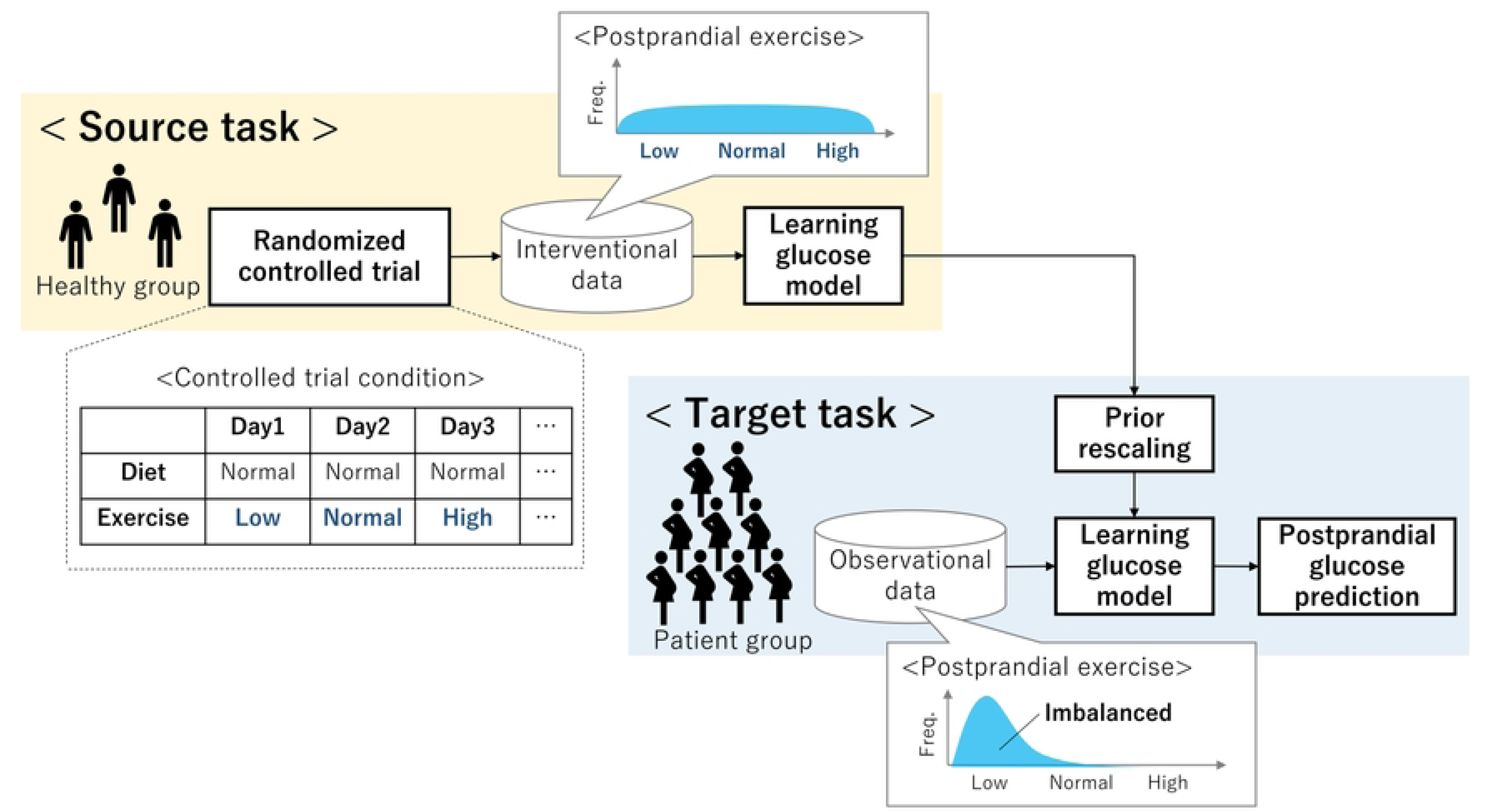
Overview of proposed transfer learning framework.

## Related work

### A. Integrative glucose prediction with diet and physical activity

Numerous machine learning techniques incorporate dietary and exercise factors to anticipate blood glucose levels. Jankovic et al. [7] and Xie [17] introduced an autoregressive model, wherein carbohydrate intake influences and muscular energy expenditure-driven exercise effects independently impact glucose levels. Likewise, support vector regression [18] and a physiological model [19] have been advanced for glucose prediction by integrating time-varying dietary and exercise influences, as computed through ordinary differential equations.

However, these methodologies entail training predictive models using individual patient historical data collected in unconstrained settings. Yet, these uncontrolled data settings introduce model misspecifications. This stems from the inherent imbalance in dietary or exercise feature distribution, attributed to each patient’s distinct and established lifestyle. Consequently, limitations in data volume per patient compound the issue.

### B. Glucose prediction with transfer learning

The primary hurdle in blood glucose level prediction lies in the scarcity of both the quality and quantity of patient data necessary for robust model training. Particularly, sophisticated deep learning techniques demand substantial training data volumes. In recent times, various strategies employing transfer learning to address this challenge have emerged. Transfer learning endeavors to construct an apt model for a target domain (referred to as a target task) by extrapolating model knowledge acquired from other domain datasets (termed source tasks) [20].

For instance, Faruqui et al. [8] suggested an approach entailing initial model learning using population data from a patient group as a source task, followed by transferring the model for individual patient-specific learning. Other studies have filtered a subset of population data based on its resemblance to a target patient, employing it as training data for either a source task [8] or a target task [15]. Furthermore, to facilitate knowledge sharing among patients, a multitasking learning strategy [21] was proposed, addressing individual model learning for all patients in parallel. Additionally, for harnessing data from diverse patient groups, adversarial transfer learning [14] was proposed, which pre-aligns feature presentations between patient groups.

Diverging from these approaches, the introduced framework (Fig 1) takes a distinct stance. Primarily, while prevailing methods seek to augment individual data volume by integrating other patient data, our approach focuses on enhancing observational data quality through leveraging data from randomized controlled trials (RCTs). Secondly, our framework stands apart by proactively intervening to procure high-quality data, with experimental conditions randomized based on the target model structure intended for learning.

## Methods

In this section, we elucidate the problem’s context, expound on the modeling of dietary and exercise impacts for postprandial glucose prediction, and subsequently detail the implementation of transfer learning utilizing the RCT dataset.

### A. Problem setting

Our model is based on an interpretable Bayesian regression model, as shown in Fig 3. We aimed to develop a predictive model that integrates the intertwined influences of both diet and exercise. Consequently, our study concentrated on forecasting postprandial glucose levels within the context of concurrent dietary and exercise effects on blood glucose. The anticipation of postprandial glucose levels assumes paramount significance in furnishing optimal behavioral suggestions, given the consistent post-meal surge in glucose levels, prone to deviations from the norm [22,23].

Consider now postprandial glucose levels with regard to the patient’s diet. When the start time of the diet is 𝜏^∗^, the target postprandial glucose level is represented as time series 𝒚_𝜏_∗+1 :𝜏∗+𝑇 of the target patient. Because the glucose level within 1 h after a diet is of clinical importance, we set T = 90 min. In addition, it is well known that carbohydrate intake (CI) increases glucose, energy expenditure (EE) from exercise lowers glucose immediately, and the features of CI [24] and EE [7] perform well for glucose prediction. Suppose, we have three types of observation variables from the patient: (i) the glucose level history before the diet 𝒚^ℋ^_1:τ∗_, (ii) the CI sequence 𝐱_1:𝑀_ from the diet and the corresponding intake timing sequence 𝐱_1:𝑀_, and (iii) the EE sequence 𝐳_1:𝑁_ from an exercise around a diet and the corresponding exercise timing sequence 𝝉_𝒛1:𝑁_. N and M denote the number of carbohydrate intakes and exercises, respectively, within one diet. Accordingly, as illustrated in Fig 2, we aim to solve a time-series regression problem to predict the target variable 𝒚_𝜏_∗+1:𝜏∗+𝑇 from observable variables 𝒚^ℋ^_1:τ∗_, 𝐱_1:𝑀_, 𝐳 1:𝑁, 𝝉 𝐱_1:𝑀_, 𝝉 𝒛1:𝑁. In the following part, these observable variables are represented without the subscript such as 𝒚,𝒚^ℋ^,𝐱, 𝐳,𝝉_𝐱_,𝝉_𝒛_ respectively.

**Fig 2.**
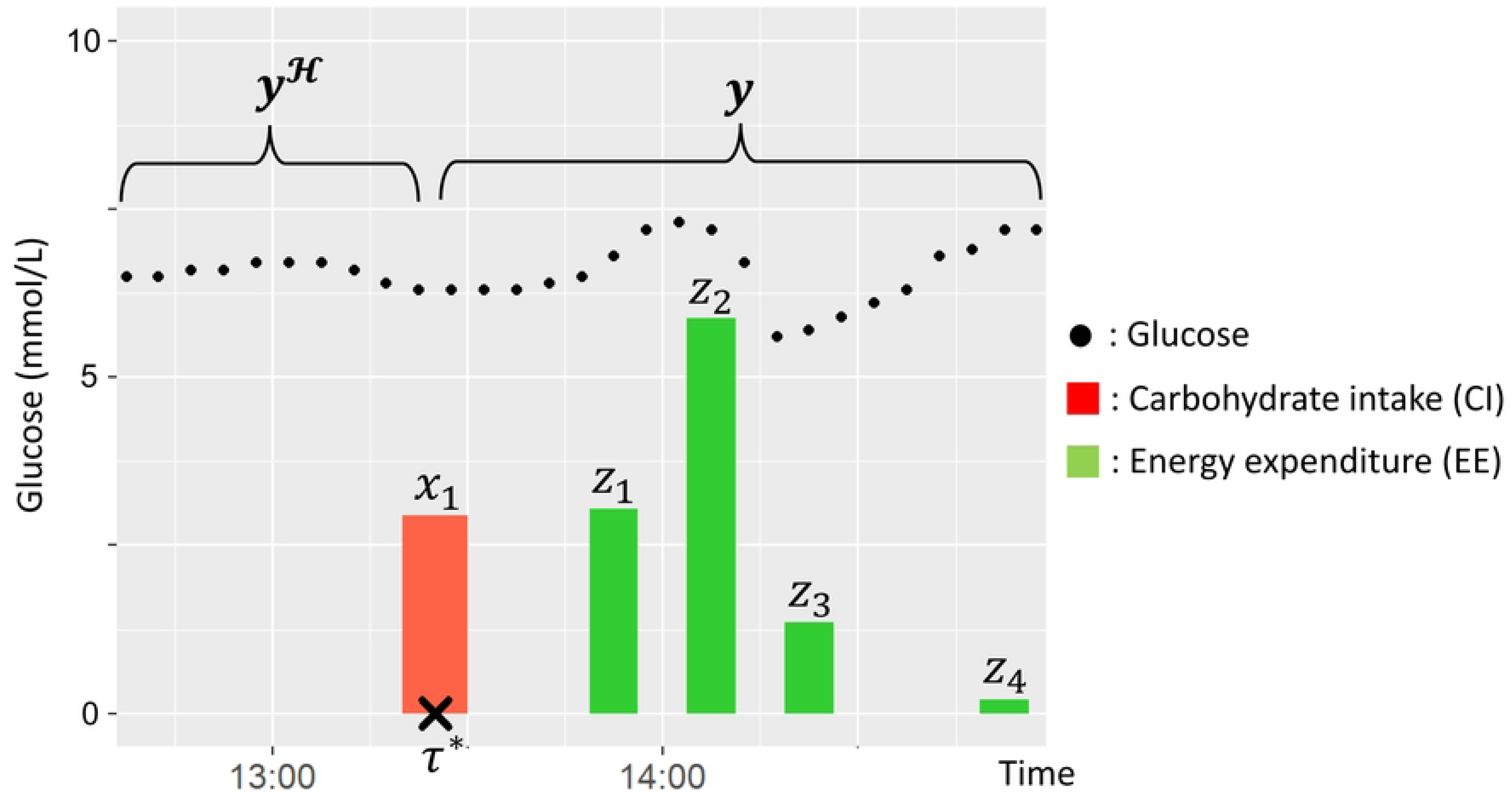
Illustration of each variable for predicting postprandial glucose.

### B. Bayesian predictive model for postprandial glucose

Our model is based on an interpretable Bayesian regression model, as shown in Fig 3. This preference is rooted in the model’s inherent transparency and traceability in contrast to complex machine learning constructs like LSTM. This transparency holds paramount significance in ensuring effective quality control for the model’s real-world applications.

**Fig 3.**
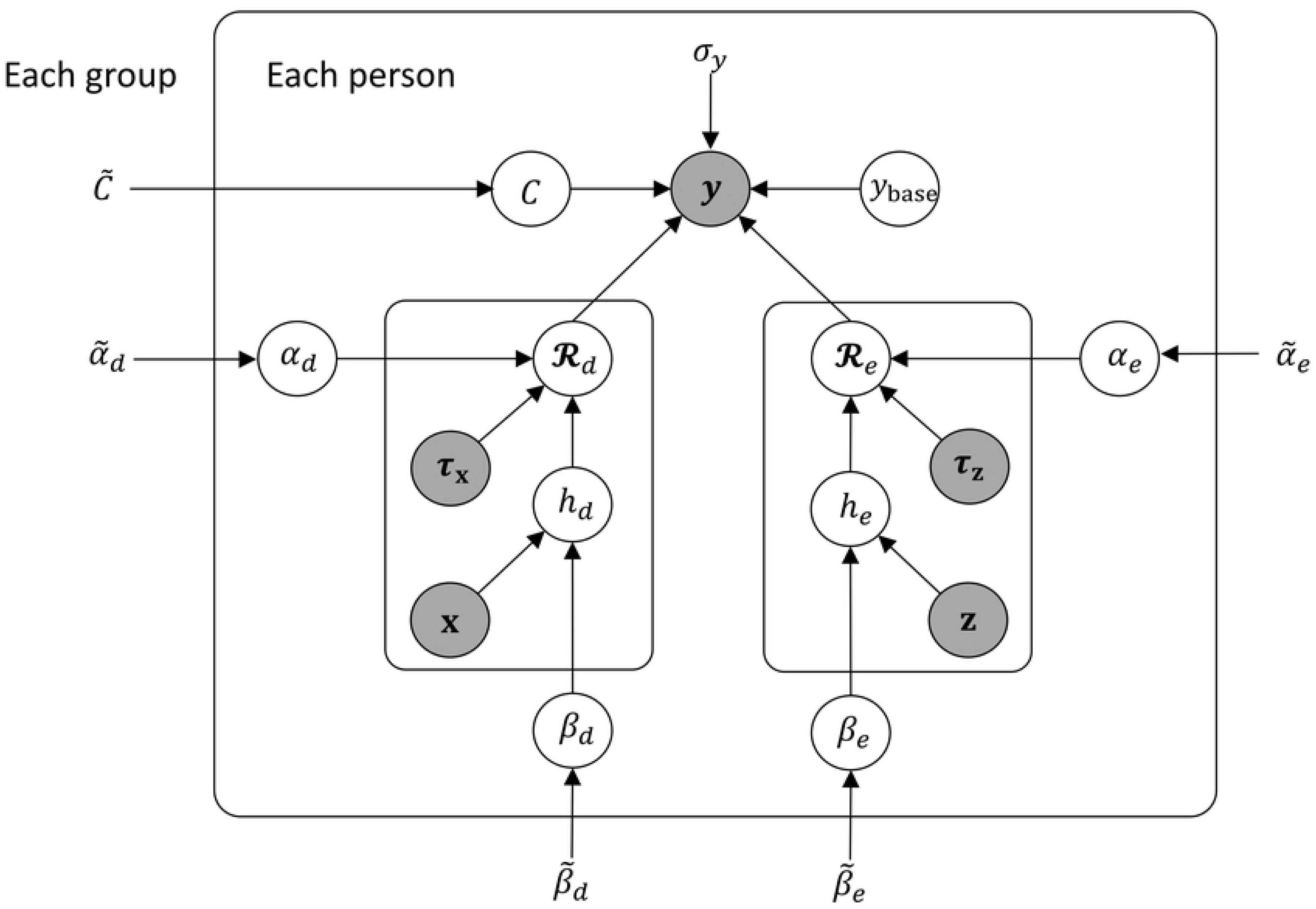
Graphical model of postprandial glucose. Parameter sets of both healthy group and patient group are estimated separately with this same model.

Following this approach, our model is based on a cutting-edge Bayesian model for glucose prediction [25], wherein forthcoming blood glucose levels are prognosticated as a summation of the time-series response under a treatment—like carbohydrate intake—and a baseline glucose level, with Gaussian noise introduced. Our study delves into two variants: an additive model and a synergistic model, both designed to account for combined effects. In the former, dietary and exercise responses are discretely generated and linearly aggregated, aligning with preceding research [7,17]. Conversely, the latter model embraces interdependency between dietary and exercise responses in a synergistic manner. This is substantiated by contemporary medical insights that highlight how the impact of postprandial exercise on glucose reduction is contingent upon carbohydrate intake levels [26] and the elevation in postprandial blood glucose [27]. Our supposition in the synergistic model postulates that such interactive impacts manifest through the multiplication of dietary and exercise effects. Furthermore, for performance benchmarking, we also explore a solitary-effect model relying solely on dietary effects, as previously addressed [25].

Specifically, the single-effect, additive-effect, and synergetic-effect models are represented by the following equations:

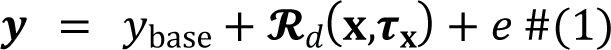

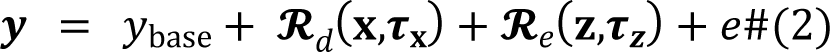

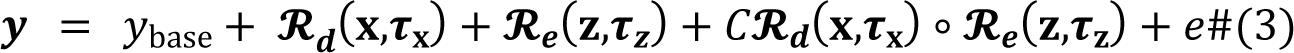

where 𝒚,𝓡_𝒅_, and 𝓡_𝒆_ are time series, and 𝑦_base_ represents the baseline blood glucose level, excluding the effects of diet and exercise. Since the time range for focusing on postprandial blood glucose is quite short, we assume that 𝑦_base_ is constant and substitute the median value of the history of preprandial blood glucose 𝒚^ℋ^from 15 min before the meal. 𝓡_𝑑_ indicates the dietary effect of increasing glucose levels, and 𝓡_𝑒_ indicates the exercise effect of decreasing glucose levels. 𝑒 represents the Gaussian noise following 𝑁(0,𝜎). Fig 3 shows a graphical representation of the synergistic model.

Furthermore, 𝓡_𝑑_and 𝓡_𝑒_are represented as time-series responses to each CI or EE treatment. Since patients occasionally have successive meals within 90 min and often perform postprandial exercise multiple times, 𝓡_𝑑_ and 𝓡_𝑒_are modeled as the summation of responses for multiple treatments in the same way as below.

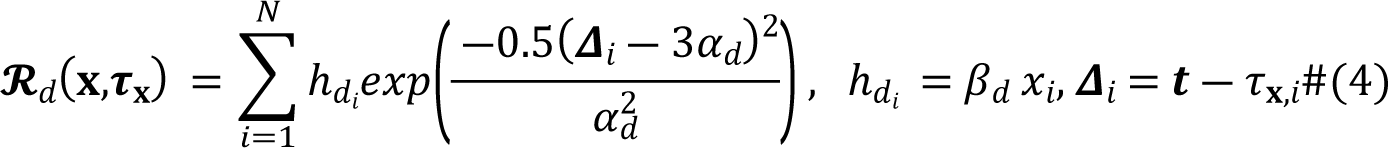

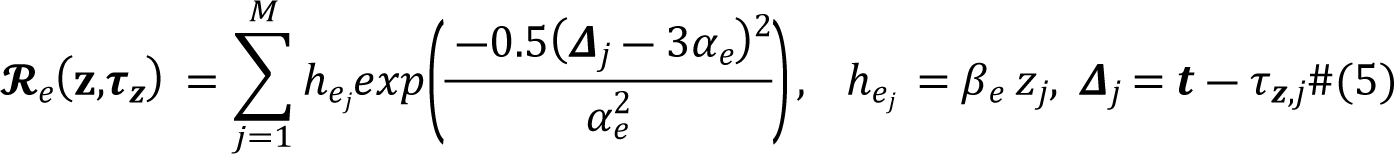

where N and M denote the numbers of meals and exercise sessions, respectively. Here, we adopt a bell-shaped function as the response function, following [25], because of its interpretability and smaller number of parameters. In this function, 𝜏_𝐱,𝑖_ and 𝜏_𝒛,𝑗_ represent the times when CI and EE start to occur, respectively. In addition, the functions are amplified by the treatment dose, that is, the amount of CI (x_𝑖_) of *i*-th intake or the amount of EE (𝑧_𝑗_) of *j*-th exercise, for each. 𝛽_𝑑_ and 𝛽_𝑒_ are parameters representing the strength of the above amplification for each treatment dose, while 𝛼_𝑑_ and 𝛼_𝑒_ are parameters representing the response speed to each treatment. Examples of 𝓡_𝑑_and 𝓡_𝑒_ are shown in Fig 4. Furthermore, to enable synergistic effect model in Eqs. 3, we introduce the adjustment parameter *C* for weighting the synergetic effect represented by 𝓡_𝑑_ ∘ 𝓡_𝑒_, where ∘ means the element-wise product.

**Fig 4.**
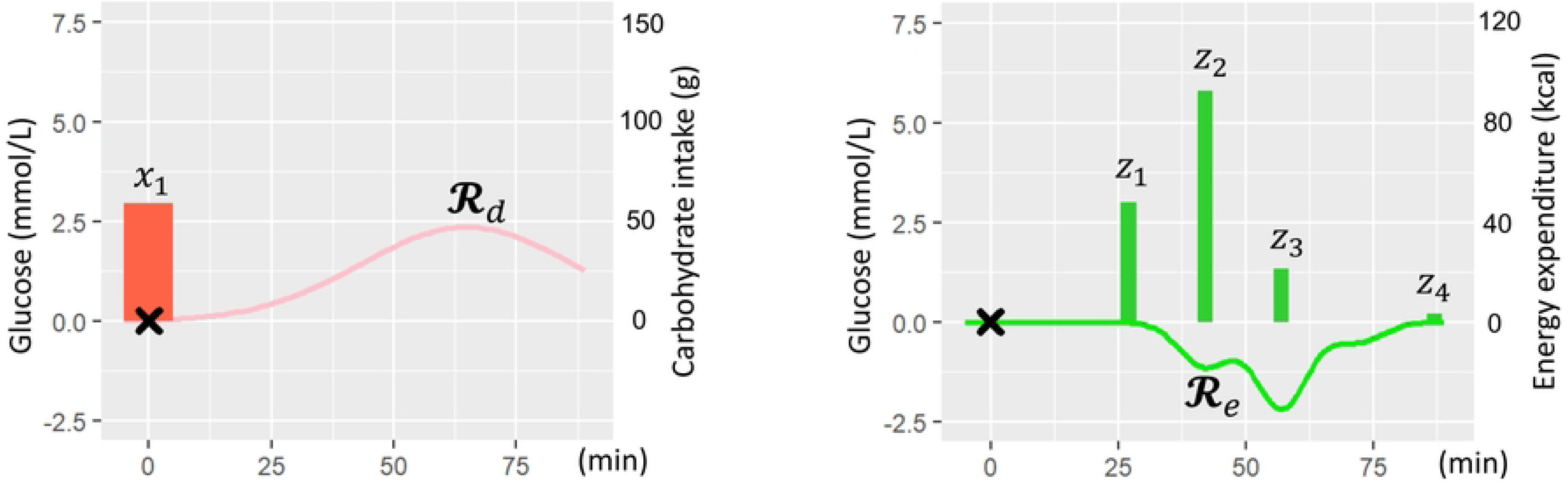
Illustration of dietary response (left) and exercise response (right) to glucose level.

Each of these parameters is patient-specific. Moreover, we introduce a hierarchical prior distribution of patient-specific parameters, enabling stable parameter learning by sharing parameter knowledge across individuals. We assume this hierarchical prior follows Gaussian centered on 𝚯̃ = (𝛼̃_𝑑_,𝛽̃_𝑑_, 𝛼̃_𝑒_,𝛽̃_𝑒_) for an additive model and 𝚯̃ = (𝛼̃_𝑑_,𝛽̃_𝑑_, 𝛼̃_𝑒_,𝛽̃_𝑒_,𝐶) for a synergetic model. These hyperparameters are common to each person in the same group (healthy or patient group) and are learned for each group, as shown in Fig 3.

### C. Bayesian transfer learning with prior rescaling from RCT data

Free-environment patient data are imbalanced in the distribution of the amount of each treatment (e.g., moderate-intensity exercise after a diet is significantly infrequent), which causes overfitting of the above hyperparameter set 𝚯̃. To address this challenge, in our proposed framework, a hyperparameter set 𝚯̃ of patient group domain is learned through transfer learning with RCT data actively collected from healthy group domain.

Initially, we direct our attention towards enlisting healthy individuals as participants for the randomized controlled trial (RCT). This choice stems from their comparative acceptability to be intervened owing to fewer underlying health issues. To achieve balanced dose distributions for each treatment, data collection is structured to randomize treatment conditions systematically. Following this, leveraging the distributional insights garnered from the acquired RCT data, we facilitate effective learning within the patient group domain by applying and adapting the knowledge embedded in the learned hyperparameter set.

In addition, our target parameters of knowledge transfer are limited to only the exercise-related hyperparameter set 𝚯̃_𝑒_ = (𝛼̃_𝑒_, 𝛽̃_𝑒_,𝐶̃) from a set of 𝚯̃, because the principle of the dietary effect to increase glucose differs between the diabetic group and the healthy group due to the significant difference in insulin sensitivity. In our RCT, we control only for the amount of exercise after the diets for each participant in the healthy group. The experimental procedure is described in the next section.

Based on the above premise, in the context of transfer learning, our source task is to learn the exercise-related hyperparameter set 𝚯̃^𝕊^_*e*_ in the healthy group domain with interventional data under RCT, and our target task is to learn the hyperparameter set 𝚯̃^𝕋^_*e*_ in the diabetic group domain with observational data under free environments.

In this study, we adopt a comprehensive framework for prior rescaling as introduced by Xuan et al. [20] in the context of Bayesian Transfer Learning (BTL). This framework entails learning the probability distribution of parameters within the source task, subsequently rescaling this learned distribution, and employing it as an informative prior within the target task, as depicted in Fig 5(b). Pertaining to this rescaling process, a technique [28] was put forth, specifically addressing the scaling of variance parameters using pre-estimated coefficients for the target task, while preserving the mean parameter. Notably, adapting this framework to our specific challenges presents intricacies. To elucidate, the influence of exercise on glucose dynamics within the healthy cohort might diverge from that within the diabetic group. Evidently, differences in the efficacy of glucose uptake in leg muscle tissues between healthy and diabetic groups emerge [29]. Consequently, this underscores the need for a mean parameter shift operation for our cross-domain prior prior to rescaling.

**Fig 5.**
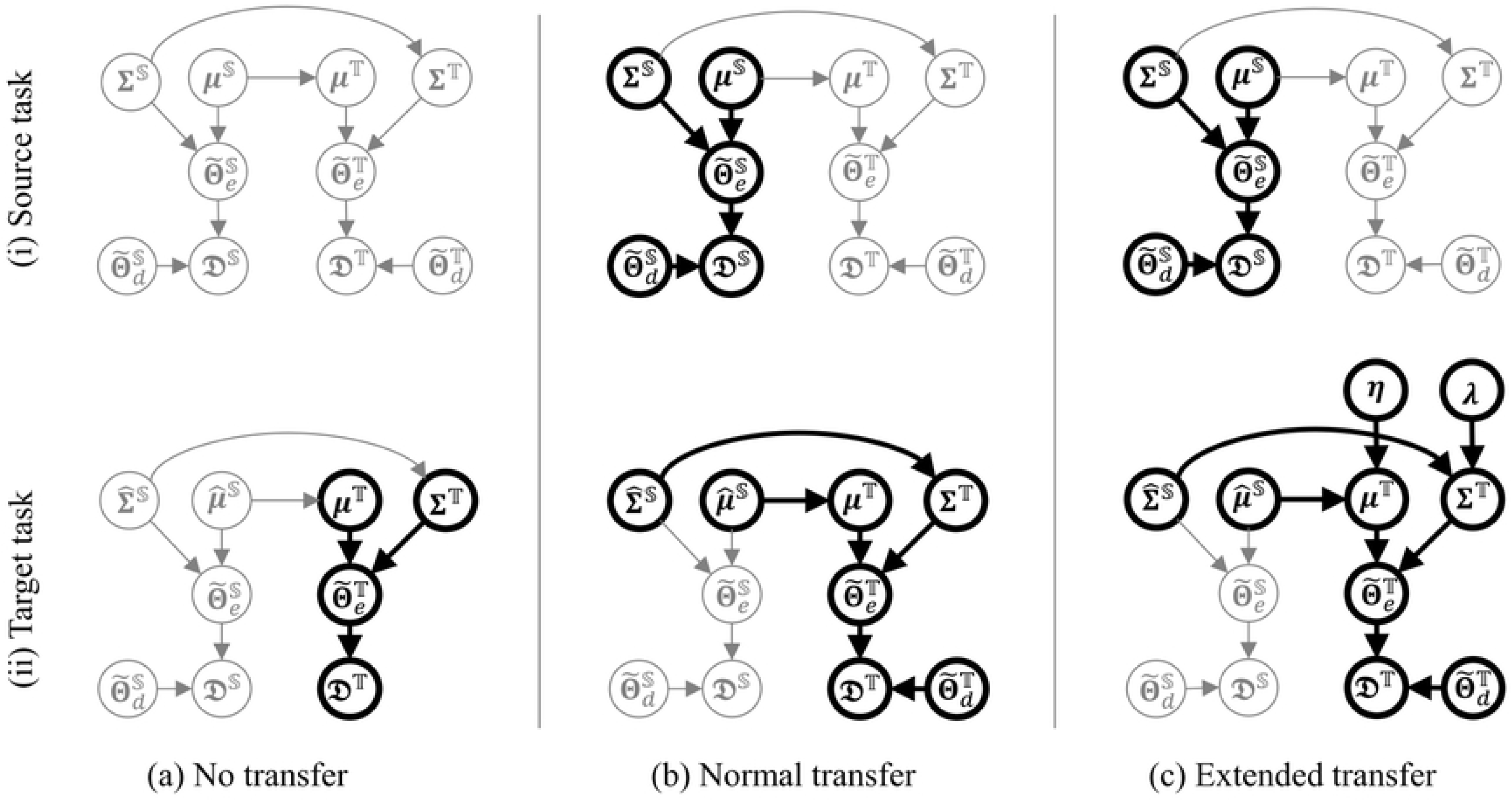
Extended framework for transfer learning. In each transfer method, only relationships between variables represented as bold line are used for parameter learning.

In this context, we propose extending the general framework to robustly shift the mean of the pretrained distribution of the parameter set 𝚯̃_𝑒_ based on clinical domain knowledge in prior rescaling, as shown in Fig 5(c). We aim to realize the distribution

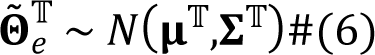

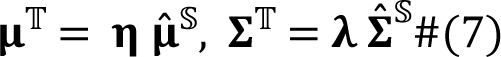

shift by introducing a new adjustment parameter 𝛈 to modulate the mean of the parameter 𝛽̃_𝑒_ representing the strength of the exercise effect on the glucose trajectory. For this, 𝜂 = 0.5 is suitable because the experimental results in the Ref. [29] show that the glucose uptake efficiency of the leg muscle in the diabetic group was about half that of the healthy group. 𝜂 for the other exercise effect parameters 𝛼̃_𝑒_ and 𝐶 in 𝚯̃_𝑒_are assumed to 1. Furthermore, another adjustment parameter 𝛌 is introduced to robustly stabilize this distribution shift in the actual training process, and this parameter 𝛌 is used to reduce the variance of each parameter. Adding this control prevents the overfitting triggered by imbalanced exercise data in the target domain. To manage the uncertainty in setting 𝛈 and 𝛌, we introduce hyperpriors for these parameters. We then use Hierarchical Bayesian estimation with the patient dataset to determine their optimal values. Based on this, prior rescaling for the target parameter set 𝚯̃^𝕋^_*e*_ in the target task is represented by the following equation:

The parameter set 𝚯̃_𝑒_ follow Gaussian distribution with a diagonal matrix 𝚺 where a variance for each parameter element is independent of each other. And 𝛈 follow Gaussian hyperprior where mean parameters correspond to 0.5 for 𝛽̃_𝑒_ and 1 for others. Also, 𝛌 follow Gaussian hyperprior with setting mean parameter to 0.1 respectively. The learning process for practical parameter learning is performed in two steps. The first step is we pre-train Gaussian parameters 𝛍^𝕊^,𝚺^𝕊^ for the distribution of 𝚯̃^𝕊^_*e*_ with RCT data in the source task. The second step is we rescale this estimand 𝛍^𝕊^, 𝚺^𝕊^ as in E.q 7, and then we perform learning all hyperparameter sets 𝚯̃^𝕋^_*e*_,𝚯̃^𝕋^_*d*_ with observational patient data, in conjunction with learning individual parameters 𝚯̃^𝕋^_*e*_,𝚯̃^𝕋^_*d*_for each diabetes patient. These parameter learnings are performed by executing a Markov Chain Monte Carlo (MCMC) simulation with the No U-Turn Sampler implemented in RStan [35].

## Experimental setup

This paper addresses two pivotal research questions.

i. How should the utilization of an RCT dataset for transfer learning be structured to acquire a predictive glucose model from an imbalanced patient dataset?
ii. How can the fusion of diet and exercise be effectively modeled to optimize the prognostication of postprandial glucose levels?

To answer these questions, we built multiple patterns of predictive models with multiple patterns of transfer learning and compared their performance based on dedicated metrics using a real-world clinical dataset of GDM patients.

### A. Evaluation policy

First, for question (i), we compared the performance of each predictive model built with and without normal or extended transfer learning described in ‘Methods B’ subsection. Second, for question (ii), we evaluated and compared the performance of the single effect model [25], the additive model, and the synergetic model described in ‘Methods A’ as a predictive model. Finally, we compared the performance of the following seven models:

- ℳ_𝑏𝑎𝑠𝑒_ : Single-effect model
- ℳ_𝑎𝑑𝑑_ : Additive model without transfer learning
- ℳ_𝑠𝑦𝑛_ : Synergistic model without transfer learning
- ℳ_𝑎𝑑𝑑+𝑡𝑟𝑎𝑛𝑠_ : additive model with normal transfer learning
- ℳ_𝑠𝑦𝑛+𝑡𝑟𝑎𝑛𝑠_ : Synergistic model with normal transfer learning
- ℳ_𝑎𝑑𝑑+𝑡𝑟𝑎𝑛𝑠_𝑒𝑥𝑡_ : additive model with extended transfer learning
- ℳ_𝑠𝑦𝑛+𝑡𝑟𝑎𝑛𝑠_𝑒𝑥𝑡_ : Synergistic model with extended transfer learning

In cases without transfer learning, we substituted a non-informative prior for the hyperparameter set 𝚯̃^𝕋^_*e*_. Notably, for any model pattern, knowledge of exercise-related parameters is still shared among all patients in learning through the hierarchical prior (‘Methods A’ subsection). Furthermore, note the additive model has limited exercise-related parameters 𝚯̃_𝑒_ = (𝛼̃_𝑒_,𝛽̃_𝑒_) which are a subset of the parameters of the synergetic model (See Eq. 2 and 5).

### B. Clinical data

The clinical data used for our performance evaluation were from a real-world free-environmental dataset of 72 patients with GDM, including continuous glucose levels, physical activity levels, and dietary records. This dataset, sourced from real-world environments, emerged from a clinical trial orchestrated by the authors [4]. This trial was granted ethical sanction by the Ethics Committee of Helsinki and the Uusimaa University Hospital District. The recruitment effort targeted patients with GDM within the gestational window of 24–28 weeks, sourced from maternity clinics in the Helsinki metropolitan area between March 10 in 2021 and December 12 in 2022. Written informed consent was obtained from all patients, and from both parents on behalf of the infant. Data acquisition occurred across 3-day intervals in monthly sessions leading up to childbirth. It’s important to note that this analysis represents a secondary examination of the eMoM GDM study [4].

Throughout each session, a continuous glucose monitoring (CGM) system (Guardian Connect System, Medtronic Ltd.) facilitated 5-minute interval glucose tracking for every patient, illustrated in Fig 6. Concurrently, data related to physical activity were collected through a wrist-worn activity tracker (Vivosmart3, Garmin International Ltd.). Energy expenditure during exercise was automatically computed via the tracker. Dietary information encompassed nutrient quantities, including carbohydrate intake (CI), ingested at each temporal juncture, sourced from patients’ manual food logs via a food tracking application developed by Helsinki University Hospital. Nutritional data integrity was fortified through nutritionist validation calls. Further information on the experimental protocol can be found in [4].

**Fig 6.**
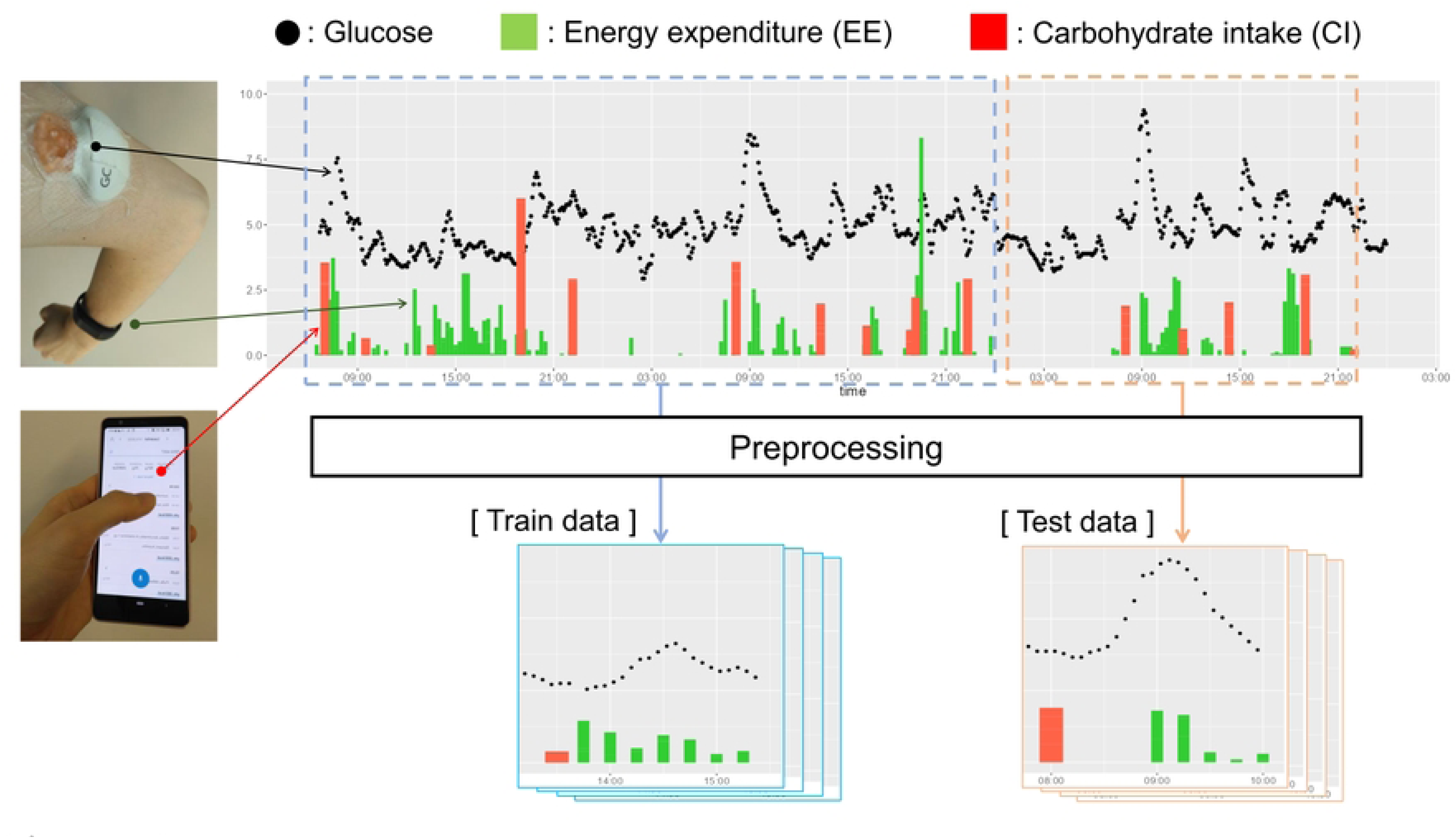
Demonstration of train and test data in a 3-days session.

### C. Preprocessing

Given our focus on postprandial blood glucose as the prediction target, we partitioned the continuous glucose data and accompanying variables around each mealtime, as depicted in Fig 6. Each segment encompassed a time span of 15 minutes preceding a meal, extending to 90 minutes post-meal. Segments with absent continuous glucose data were omitted from analysis, leading to the exclusion of 4 patient datasets. As a result, 1619 segment of data were obtained from 68 patients. Subsequently, the segment data in the first two days of each session were used as the training data 𝓓^𝕋^_*train*_, and the segment data 𝑡𝑟𝑎𝑖𝑛 in the last day were used as the test data 𝓓^𝕋^_*test*_. The predictive model was trained in each session and its evaluation was 𝑡𝑒𝑠𝑡 performed within that session. This is because the segment data of the next month’s session are heterogeneous from that of the current month, as the condition of a pregnant woman changes drastically after one month, even for the same individual [39].

### D. Randomized controlled trial

The RCT data for our source task were collected from additionally recruited 4 healthy subjects (Aalto University students) in a six-days session with the same data collection as that in the clinical trial. The recruitment period was from February 1 in 2023 to April 30 in 2023. Written informed consent was obtained from all the participants for data collection and utilization. The purpose of the RCT was to obtain a segmented dataset in which the distribution of EE during postprandial exercise enables robust learning of the exercise-related parameter set 𝚯̃^𝕊^. Therefore, the postprandial exercise conditions during data collection were randomized for each participant. In practice, the subjects were instructed to follow different conditions, as follows (See Fig 1).

- **Low-intensity condition (Day 1 & Day 4)**

♣ Eat almost same amount of carbohydrate at lunch (or dinner)
♣ Don’t perform an exercise until two hours after eating.
- **Moderate-intensity condition (Day 2 & Day 5)**

♣ Eat almost same amount of carbohydrate at lunch (or dinner)
♣ 30 minutes after starting eating, walk continuously at a pace of 100 step/min for 20 minutes.
♣ After walking, don’t perform an exercise until two hours after eating.
- **High-intensity condition (Day 3 & Day 6)**

♣ Eat almost same amount of carbohydrate at lunch (or dinner)
♣ 30 minutes after starting eating, walk continuously at a pace of 130 step/min for 20 minutes.
♣ After walking, don’t perform an exercise until two hours after eating.

The above exercise conditions were designed based on clinical findings of the effect of exercise on glucose. Specifically, according to the literature [30], the appropriate timing of exercise for decreasing blood glucose was reported to be 30 min after the start of meals, when the exercise content was continuous walking for 20 min.

While the exercise condition differed between the days, the dietary condition was controlled to be the same for all days for each subject, as shown in Fig 1. This is because by equalizing the dietary effect on postprandial glucose among all days in a subject, the learning of targeted exercise parameter sets 𝚯̃^𝕊^can be facilitated more in our source task. Additionally, the participants were asked to choose breakfast or lunch as the target diet for each day.

The RCT and data preprocessing resulted in 24 segments of data 𝓓^𝕊^_*train*_. Fig 7 shows the actual distribution of the EE in the 𝑡𝑟𝑎𝑖𝑛 interventional RCT and observational GDM datasets. This confirms lesser imbalance in the distribution from the RCT dataset compared to that from the GDM dataset.

**Fig 7.**
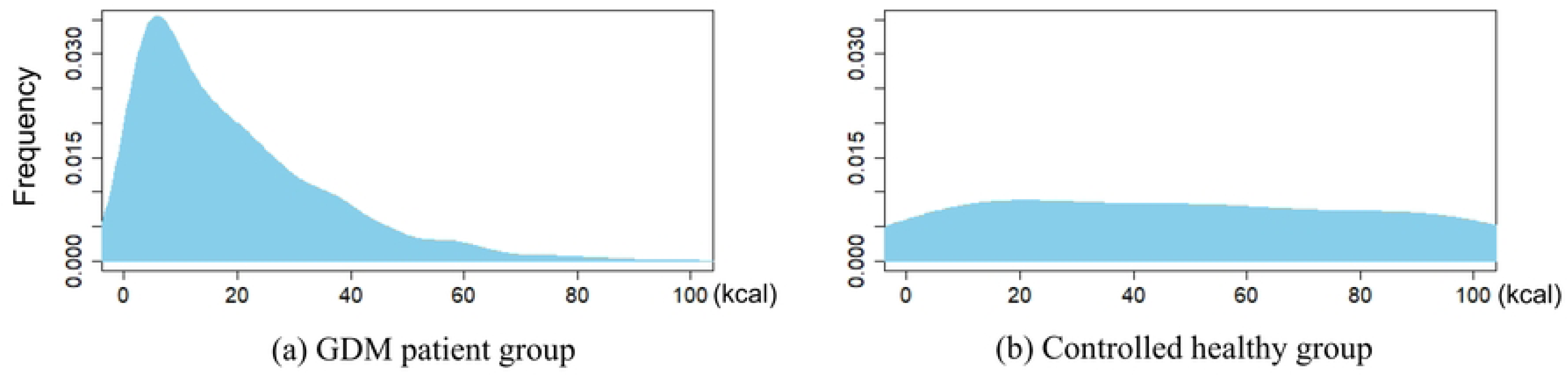
Actual distribution of energy expenditure (EE) in postprandial exercise.

### E. Parameter learning and prediction

The posterior of the overall parameter sets involved in each model among the seven patterns was estimated by MCMC simulation using both the observational training data 𝓓^𝕋^_*train*_ of the GDM group and the interventional RCT data 𝓓^𝕊^_*train*_ of the healthy group. Subsequently, we obtained the point estimation values 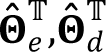 which are a set of a posteriori medians for each 𝑒 𝑑 parameter for each patient. Then, the predicted future postprandial glucose trajectory 𝒚_𝜏_∗+1:𝜏∗+𝑇 was obtained by applying the model embedded with 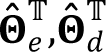 to the observed variable values 𝒚^ℋ^,𝐱,𝒛,𝝉, 𝝉 in the test data 𝓓^𝕋^_*test*_ of the GDM group.

### F. Metrics

The most important evaluation criterion is the extent to which the predicted glucose series 𝒚 is coincident with the actual observation series 𝒚. Therefore, we evaluated (1) the root mean squared error (RMSE) of the predicted series and (2) the mean absolute error (MAE). Additionally, we evaluated the degree of coincidence of (3) the area under the curve (AUC) and (4) the postprandial maximum value of the glucose series because these are well-known glycemic indicators for diabetes research [36].

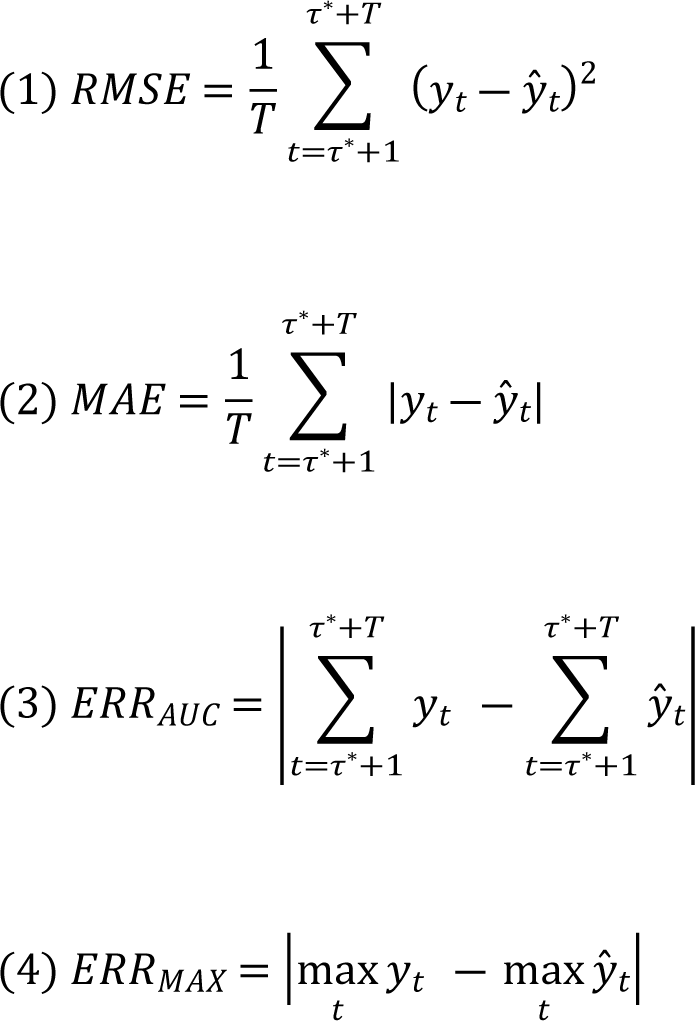

After calculating each metric following the above equations for all segment data included in the test data 𝓓^𝕋^_*test*_, the average of the metric values among all segments was used as the final metric score.

## Result

Table 1 presents the conclusive average metric scores for each model across segments both with and without postprandial exercise. Within this context, an exercise segment is delineated by an energy expenditure (EE) exceeding 60 kcal. Focusing initially on segments involving postprandial exercise, we observe that the synergetic model, trained through extended transfer learning with RCT data, achieves the highest performance. Additionally, the augmentation of performance is evident across both the additive and synergetic models due to extended transfer learning, affirming its efficacy. Notably, this enhancement is particularly pronounced in the synergetic model, attributed to its more intricate model structure. Conversely, across segments devoid of postprandial exercise, the metrics remain largely consistent across all models. This consistency aligns with expectations, given that the exercise effect (*R_e_*) within the additive or synergetic model approximates zero in the absence of postprandial exercise. Consequently, the forecasted glucose trajectory converges with that of the single model (Eq. 1-3).

**Table 1.**
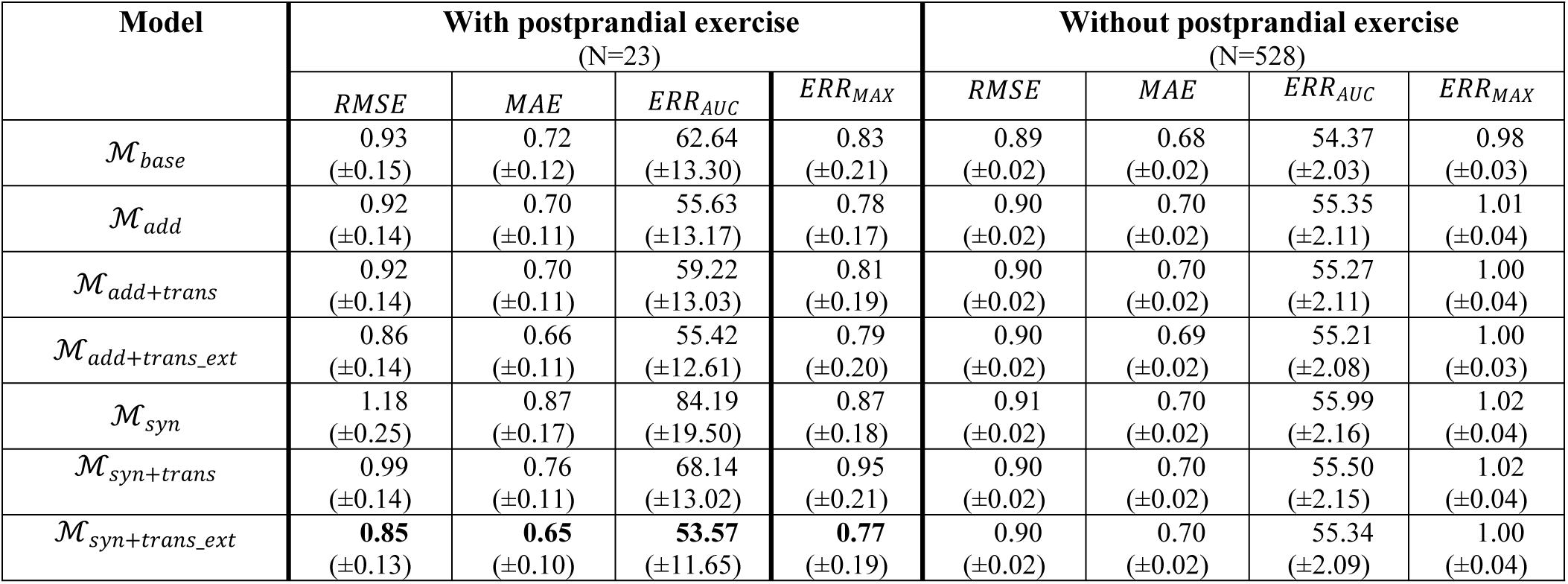
Average metric scores with standard error.

| Model | With postprandial exercise<br>(N=23) |  |  |  | Without postprandial exercise<br>(N=528) |  |  |  |
| --- | --- | --- | --- | --- | --- | --- | --- | --- |
|  | *RMSE* | *MAE* | *ERR<sub>AUC</sub>* | *ERR<sub>MAX</sub>* | *RMSE* | *MAE* | *ERR<sub>AUC</sub>* | *ERR<sub>MAX</sub>* |
| $\mathcal{M}_{base}$ | 0.93<br>($\pm 0.15$) | 0.72<br>($\pm 0.12$) | 62.64<br>($\pm 13.30$) | 0.83<br>($\pm 0.21$) | 0.89<br>($\pm 0.02$) | 0.68<br>($\pm 0.02$) | 54.37<br>($\pm 2.03$) | 0.98<br>($\pm 0.03$) |
| $\mathcal{M}_{add}$ | 0.92<br>($\pm 0.14$) | 0.70<br>($\pm 0.11$) | 55.63<br>($\pm 13.17$) | 0.78<br>($\pm 0.17$) | 0.90<br>($\pm 0.02$) | 0.70<br>($\pm 0.02$) | 55.35<br>($\pm 2.11$) | 1.01<br>($\pm 0.04$) |
| $\mathcal{M}_{add+trans}$ | 0.92<br>($\pm 0.14$) | 0.70<br>($\pm 0.11$) | 59.22<br>($\pm 13.03$) | 0.81<br>($\pm 0.19$) | 0.90<br>($\pm 0.02$) | 0.70<br>($\pm 0.02$) | 55.27<br>($\pm 2.11$) | 1.00<br>($\pm 0.04$) |
| $\mathcal{M}_{add+trans\_ext}$ | 0.86<br>($\pm 0.14$) | 0.66<br>($\pm 0.11$) | 55.42<br>($\pm 12.61$) | 0.79<br>($\pm 0.20$) | 0.90<br>($\pm 0.02$) | 0.69<br>($\pm 0.02$) | 55.21<br>($\pm 2.08$) | 1.00<br>($\pm 0.03$) |
| $\mathcal{M}_{syn}$ | 1.18<br>($\pm 0.25$) | 0.87<br>($\pm 0.17$) | 84.19<br>($\pm 19.50$) | 0.87<br>($\pm 0.18$) | 0.91<br>($\pm 0.02$) | 0.70<br>($\pm 0.02$) | 55.99<br>($\pm 2.16$) | 1.02<br>($\pm 0.04$) |
| $\mathcal{M}_{syn+trans}$ | 0.99<br>($\pm 0.14$) | 0.76<br>($\pm 0.11$) | 68.14<br>($\pm 13.02$) | 0.95<br>($\pm 0.21$) | 0.90<br>($\pm 0.02$) | 0.70<br>($\pm 0.02$) | 55.50<br>($\pm 2.15$) | 1.02<br>($\pm 0.04$) |
| $\mathcal{M}_{syn+trans\_ext}$ | **0.85**<br>($\pm 0.13$) | **0.65**<br>($\pm 0.10$) | **53.57**<br>($\pm 11.65$) | **0.77**<br>($\pm 0.19$) | 0.90<br>($\pm 0.02$) | 0.70<br>($\pm 0.02$) | 55.34<br>($\pm 2.09$) | 1.00<br>($\pm 0.04$) |

The variations in the projected glucose trajectory by the synergetic model for each training scheme are depicted in Fig 8. Instances of (a) no transfer and (b) regular transfer reveal discrepancies between the projected value (dark pink line) and the actual value (black dot). This discord stems from an overestimation of the exercise effect, which hampers the glucose response induced by diet (light pink line) subsequent to exercise events (green bar). In contrast, Fig 8(c) showcases the efficacy of extended transfer learning, encompassing a distributional shift from the RCT dataset. This approach ensures an appropriately calibrated exercise effect in terms of intensity and timing, consequently yielding ameliorated prediction errors.

**Fig 8.**
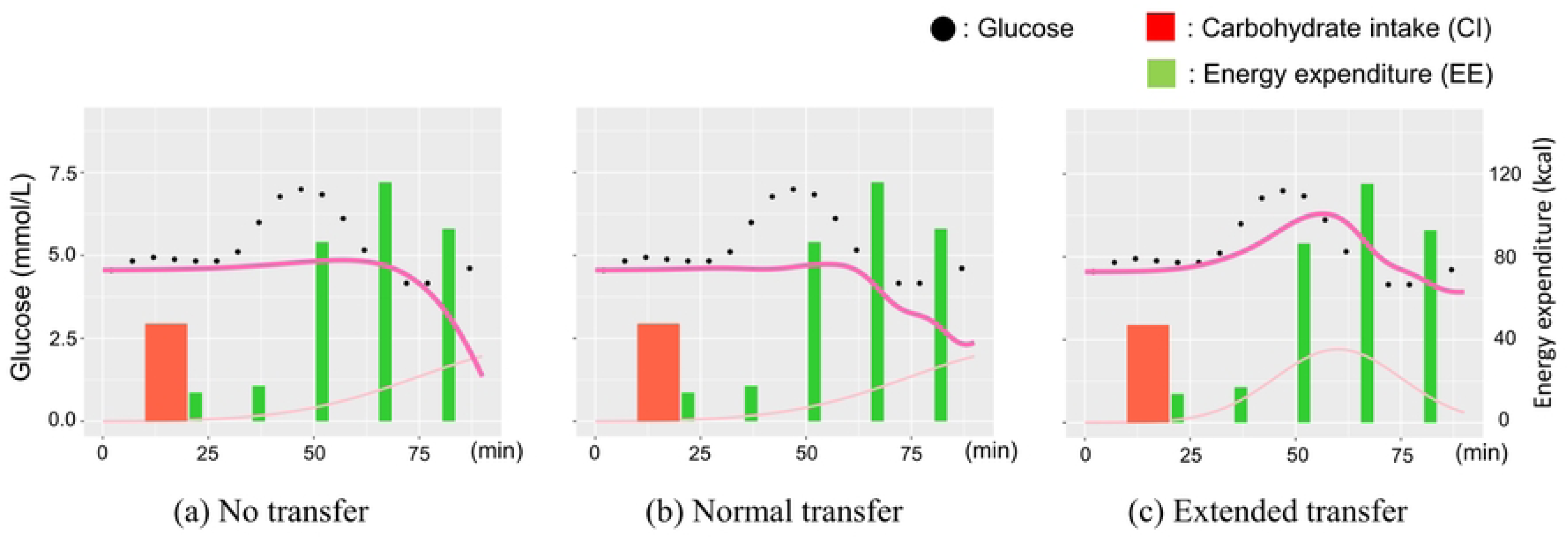
Difference in predicted glucose trajectory with each training pattern.

Furthermore, Fig 9 illustrates instances of projected trajectories by the single, additive, and synergetic models. Notably, the glucose surge projected by the single model (a) exhibits a pronounced delay compared to the actual rise. This delay is likely attributed to overfitting of dietary parameters due to the omission of the exercise effect. Contrastingly, the combined models (b) and (c) aptly replicate the glucose elevation, demonstrating their success in addressing this aspect.

**Fig 9.**
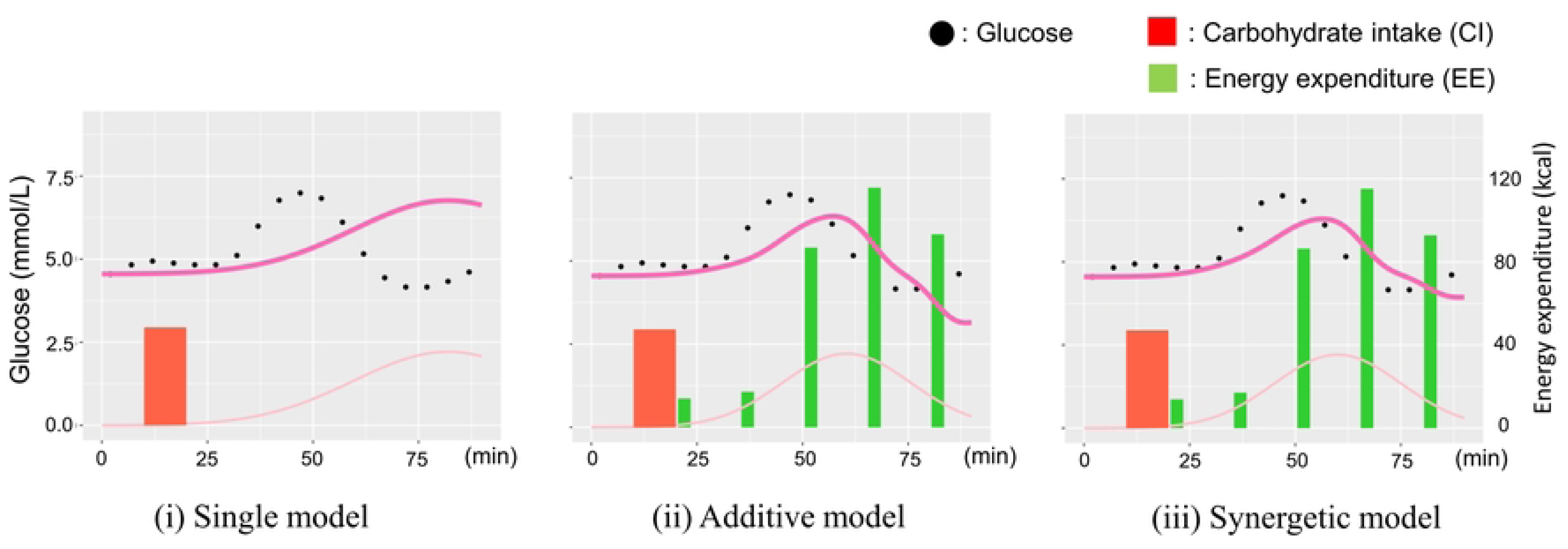
Examples of trajectory predicted by each glucose model.

These results demonstrate the effectiveness of our transfer learning framework with RCT data and the synergistic modelling of dietary and exercise effects on glucose.

## Discussion

An intrinsic strength of the proposed methodology lies in the transparency and visibility of the trained models and their parameters, as depicted in Fig 3. This transparency facilitates seamless incorporation into the formulation of personalized behavioral recommendations for individual patients. Moreover, the integration of transfer learning with randomized controlled trial (RCT) data notably amplifies this capability by refining model parameters with heightened precision. For example, if the absolute value of exercise parameter 𝛽̃_𝑒_ in Eq. Five is estimated to be small in some patients, the exercise effect 𝓡_𝑒_ is not likely to appear easily, implying that recommendations regarding postprandial exercise should be of moderate (or higher) intensity. Here, Fig 10 shows actual examples of the posterior of the parameter 𝛽̃_𝑒_ estimated for each healthy participant on the top and each patient on the bottom. As the example in Fig 8 belongs to patient P1 in Fig 10, we can confirm the overestimation of 𝛽̃_𝑒_ from Fig 10(a) and (b) at the bottom. In the context of practical recommendation scenarios, this tendency results in excessively optimistic and potentially detrimental suggestions, assuming a minor exercise could significantly enhance the patient’s glucose profile. Yet, as depicted in Fig 10(c), this misalignment is rectified through extended transfer learning, facilitated by prior rescaling via the incorporation of shifting and shrinking operations outlined in Eq. 7. This underscores the efficacy of the proposed transfer learning technique, enabling the formulation of exercise recommendations that genuinely optimize postprandial glucose reduction for individual patients, thanks to the precise acquisition of parameters.

**Fig 10.**
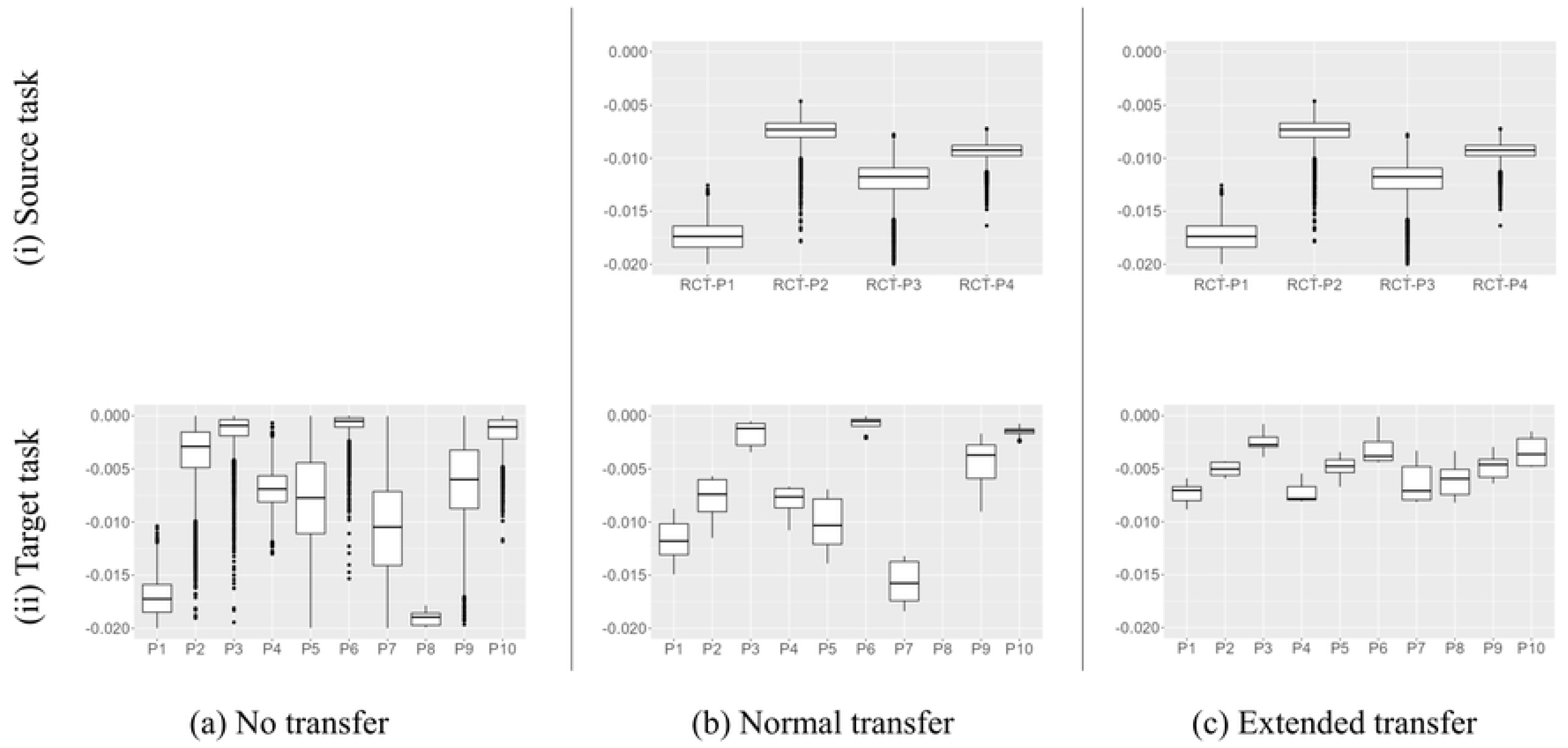
Estimated distribution of parameter **𝛽̃_𝑒_** for each patient.

An additional advantage offered by the proposed transfer learning technique is its intrinsic ability to automatically establish a fitting prior distribution. Practical Bayesian modeling often entails substantial reliance on domain-specific knowledge for setting informative priors [31]. Nevertheless, acquiring such requisite knowledge for training intricate time-series models is notably challenging, given the nascent state of corresponding medical insights in numerous cases [32]. In response to this challenge, our approach empowers the creation of prior distributions with minimal domain knowledge, achieved through leveraging RCT data acquired via active intervention. Furthermore, due to the inherent simplicity of our devised framework (Fig 1), its applicability extends to glucose prediction for diverse patient cohorts and other complex prognostication tasks within the healthcare domain.

However, an intriguing and contentious issue revolves around determining the requisite volume of data for a dedicated RCT within our framework. Guided by the Law of Large Numbers, the greater the volume of RCT data amassed, the more balanced the target distribution becomes, consequently bolstering the robustness of parameter learning. Nevertheless, amassing a substantial quantity of high-quality RCT data necessitates significant time and resources, given the demand for large-scale experimental endeavors.

Therefore, in practice, the amount of RCT data should be determined flexibly depending on the results of the convergence diagnosis in parameter learning. Here, the values of convergence indicator 𝑅 [31] for each parameter in 𝚯̃^𝕊^ = (𝛼̃_𝑑_,𝛽̃_𝑑_,𝛼̃_𝑒_,𝛽̃_𝑒_,𝐶) were (1.0001, 1.0040, 1.0017, 1.0008, 1.0013) for each, in our source task. Because 𝑅 < 1.1 is conventionally considered that parameter learning converges, and the number of RCT could be sufficient to specify the posterior distribution of the parameters.

In our exploration of exercise’s impact, our focus thus far has centered on the immediate reduction of glucose levels owing to muscular fatigue. However, insights gleaned from prior medical investigations [33] underscore a persistent, longer-term influence of exercise on enhancing insulin sensitivity and adeptly regulating blood glucose responses through repeated vigorous physical activity in patients’ daily routines. In forthcoming endeavors, we aim to integrate these enduring exercise effects into our glucose prediction model, striving for optimal predictive accuracy and the formulation of tailored behavioral recommendations for patients. It’s worth noting that other variables such as stress, sleep patterns, time zone disparities, dietary history, and medical conditions also exert influence on glucose trajectories [6,23,37]. To refine our predictions and model training, we intend to encompass these additional factors within our glucose modeling framework in future research.

## Conclusion

We present an innovative transfer-learning framework utilizing randomized controlled trial (RCT) data for postprandial glucose prediction, integrating both dietary and exercise behaviors. The effectiveness of this framework was assessed using real-world data collected from 68 patients with gestational diabetes mellitus (GDM) in their everyday settings. The evaluation conclusively demonstrates performance enhancement in postprandial glucose prediction through the implementation of our proposed transfer learning approach. Our findings also underscore the superior accuracy of the synergetic model compared to the additive model in modeling combined factors. Moving forward, our research aims to incorporate the prolonged glycemic impact of exercise routines to forge a superior predictive model for tailored recommendations. Additionally, we will extend the application of this framework to various prediction tasks to gauge its adaptability and versatility in future.

## Data Availability

Data cannot be shared publicly because of the terms of personal information protection in our trials. Data are available from the HUS Institutional Data Access / Ethics Committee (contact via Saila Koivosalo) for researchers who meet the criteria for access to confidential data.

## Acknowledgments

The authors would like to thank Çağlar Hızlı and Arina Odnoblyudova for useful technical discussions. We also would like to thank Editage for English language editing.

